# Genome evolution and early introductions of the SARS-CoV-2 Omicron Variant in Mexico

**DOI:** 10.1101/2022.07.13.22277603

**Authors:** Hugo G. Castelán-Sánchez, León P. Martínez-Castilla, Gustavo Sganzerla-Martínez, Jesús Torres-Flores, Gamaliel López-Leal

## Abstract

A new variant of SARS-CoV-2 Omicron (Pango lineage designation B.1.1.529), was first reported to the World Health Organization (WHO) by South African health authorities on November 24, 2021. The Omicron variant possesses numerous mutations associated with increased transmissibility and immune escape properties. In November 2021, Mexican authorities reported Omicron’s presence in the country. In this study, we infer the first introductory events of Omicron and the impact that human mobility can have on the spread of the virus. We also evaluated the adaptive evolutionary processes in Mexican SARS-CoV-2 genomes during the first month of circulation of Omicron.

We infer 173 introduction events of Omicron in Mexico in the first two months of detection; subsequently, of the introductions, there was an increase in the prevalence for January.

This higher prevalence of the novel variant results in a peak of cases reported, on average, six weeks after a higher mobility trend was reported. The peak of cases reported is due to the BA.1.1 Omicron sub-lineage dominated, followed by BA.1 and BA.15 sub-lineages in the country from January to February 2022.

Additionally, we identified the presence of diversifying natural selection in the genomes of Omicron and found mainly five non-synonymous mutations in the RDB domain of the Spike protein, all of them related to evasion of the immune response. In contrast, the other proteins in the genome are highly conserved—however, there are homoplasies mutations in non-structural proteins, indicating a parallel evolution.

## 1. INTRODUCTION

The new coronavirus SARS-CoV-2, the causative agent of coronavirus disease 2019 (COVID-19), emerged at the end of 2019 in the province of Wuhan, China, and rapidly spread to other countries, causing more than 5.2 million deaths worldwide as of November 18, 2022 (Platto, Xue, y Carafoli 2020; Zhu et al. 2020). As new variants of SARS-CoV-2 emerged in different regions of the world, the World Health Organization (WHO) designated new variants as variants of interest (VOI), variants under monitoring (VUM), and variants of concern (VOC), depending on their epidemiological and clinical behavior (Aleem, Akbar Samad, y Slenker 2022). The emergence of VOCs has been of particular interest because they are associated with remarkable increases in the incidence of COVID-19 worldwide (Choi y Smith 2021). For example, the Alpha (B.1.1.7), Gamma (P.1.X), and Delta (B.1.617.2) variants were associated with drastic increases in the number of infections and COVID-19-related deaths in the United Kingdom, Brazil, and India, respectively (Tao et al. 2021).

On November 24, 2021, health authorities in Botswana and South Africa reported circulating a new SARS-CoV-2 variant that was later designated Omicron and classified as a VOC by the WHO (Viana et al. 2022). This new variant displayed several mutations across the genome, some previously found in other VOCs (Thakur y Ratho 2022). One of the most critical traits of Omicron is the presence of a high number of mutations (32 mutations) in the spike protein compared to other VOCs, which might confer this variant an increased affinity towards the angiotensin-converting enzyme 2(ACE2 receptor), enhancing entry into the host cell, and the ability to escape neutralization by antibodies induced by either natural infection or vaccination (Thakur y Ratho 2022). Moreover, recent *in vitro* studies reported that the replication rates for Omicron are almost 70 times higher in bronchial tissue than in lung tissue in contrast to the Delta variant, which might explain the high transmissibility of Omicron (Chi-wai MC, Nicholls J, Pui-yan KH, Peiris M, Wah-Ching T, Lit-man LP s/f) Omicron has already spread across 77 countries, and their genomes are available in GISAID (Thakur y Ratho 2022). The Omicron variant was first detected in Mexico in November 2021 and has rapidly spread since, probably causing the recent increase in symptomatic COVID-19 cases observed in the country. Due to the high infectivity of Omicron in contrast to other VOCs, the study of the evolution and transmission patterns of this variant in different countries is crucial for the implementation of effective strategies to reduce its impact in a clinical context. In this work, we focus on identifying the first events of Omicron introduction to understand how the virus spreads and spreads in Mexico, with which it is possible to identify more than 100 multiple independent introductions. In addition, we analyzed adaptation signals throughout the genome and found only evidence of natural selection in protein Spike and homoplasic mutations in nonstructural proteins.

## 2. MATERIALS AND METHODS

### 2.1 Collection of genomes

Two hundred fifty-four complete Omicron genomes reported in Mexico were downloaded between November 28 to December 27, 2021, from the Global Initiative on Sharing All Influenza Data GISAID platform (https://www.gisaid.org/). Additionally, the first 20 genomes of Omicron reported per country were downloaded. However, there were fewer genomes available for some because there is no comprehensive sequencing coverage. Finally, 2,148 Omicron SARS-COV-2 genomes sequenced worldwide were analyzed (including 187 of other lineages to guide the phylogeny). Additionally, to analyze the spread and diversification of Omicron in Mexico, genomes of this GISAID variant were downloaded from November 2021 to April 2022 with the parameters of inclusion, complete genome, high coverage, and low coverage exclusion.

### 2.2 Phylogenomics and phylogeographic analysis

Genomes were aligned using MAFFTv7 software (Katoh, Rozewicki, y Yamada 2019), trimming the 5’ and 3’ ends, and classified according to their viral lineage assignment under the Pango nomenclature system using Pangolin v3.1.7, version (Rambaut et al. 2020).

To estimate the phylogenetic relationships and phylogeography between SARS-COV-2 genomes, a maximum likelihood (ML) phylogenetic tree was constructed using the IQ-Tree (Nguyen et al. 2015) under the GTR + G model of nucleotide substitution with empirical base frequencies and four free site rate categories and statistical support (-alrt 1000). Sequences from lineages other than Omicron were added to this phylogeny to establish the monophyly of Omicron subtrees, while the sequences from Wuhan were used to root the tree. The tree was evaluated for a temporal signal using TempEst (Rambaut et al. 2016) (the outliers were eliminated).

A time-scaled phylogenetic tree was built with TreeTime v0.7.4 (Sagulenko, Puller, y Neher 2018), which specified a clock rate of 8×10^−4^ substitutions per site per year (s/s/y) according to the Nextstrain workflow (Hadfield et al. 2018).

The phylogeographic analysis detected and quantified Omicron lineage introduction events into Mexico. This analysis was reconstructed from the time-scaled tree generated previously with TreeTime v0.7.4. We use two different approaches based on several models and inference methods. To identify independent introduction events of Omicron in Mexico, we used a discrete diffusion model implemented in the software package BEAST v1.10.4 (Drummond et al. 2012). The Bayesian analysis through Markov chain Monte Carlo (MCMC) was run on 106 generations and sampled every 1,000 generations using the BEAST v1.10.4 tool. The time-scaled phylogeny previously built was a fixed empirical tree and considered two possible ancestral locations: “Mexico” and “other location,” according to the described pipeline Dellicour et al., 2021 (Dellicour, Durkin, et al. 2021, 2).

The MCMC convergence and mixing properties were inspected using Tracer v1.72 (Rambaut, A., Suchard, M.A., Xie, D. and Drummond, A.J. 2014, 6), and the effective sample size (ESS) values above 200 were achieved for all parameters. The phylogenies were annotated with a 10% burn-in by Tree Annotator. Subsequently, the “Seraphim” package (Dellicour et al. 2016) extracted the Spatio-temporal information from the data and visualized the phylogeographic reconstructions.

Second, we use PASTML (Ishikawa et al. 2019) briefly reconstructs the ancestral character states and their changes along with the trees, with maximum likelihood marginal posterior probabilities approximation (MPPA) and Felsenstein 1981 (F81) model options. For the parameters in PASTML, we used MPPA as a prediction method (standard settings). We added the character predicted by the joint reconstruction, even if it was not selected by the Brier score (option forced_joint).

On the other hand, the Omicron sequences from Mexico that were downloaded from November to April were built in to reconstruct a maximum likelihood phylogeny using iqtree v.2.1.1, with the GTR+F+R3 substitution model and statistical support (-alrt 1000) and the phylogenetic tree was visualized in iTOL v5 (Letunic y Bork 2021).

### 2.3 Daily mobility and cases report

The mobility data used in this study was obtained from the Google Mobility Reports service (https://www.google.com/covid19/mobility/) for all Mexican states. The data is found in the following categories: *i*) grocery and pharmacy; *ii*) retail and recreation; *iii*) parks; *iv*) workplaces; *v*) residential; *vi*) transit stations. The service has been updated daily since 26-02-2020. The last date of collection was 17-04-2022. The data were treated in weekly variations. Finally, we added up all six instances of mobility to come up with a single unitary value representing the mobility of a given week.

The country was divided into seven regions to analyze the mobility as follows: Northeast (NE; Coahuila, Nuevo León, and Tamaulipas), Northwest (NW; Baja California, Baja California Sur, Chihuahua, Durango, Sonora, and Sinaloa) Central North (CN; Aguascalientes, Guanajuato, Querétaro, San Luis Potosí, and Zacatecas), Central South (CS; Mexico City, Estado de México, Morelos, Hidalgo, Puebla, and Tlaxcala), West (W; Colima, Jalisco, Michoacán, and Nayarit), Southeast (SE; Guerrero, Oaxaca, Chiapas, Veracruz, and Tabasco) and South (S; Campeche, Yucatán, and Quintana Roo).

Moreover, daily cases of the COVID-19 transmission in Mexico were retrieved from the service https://datos.covid-19.conacyt.mx/. The regionalized (please see above) daily cases were analyzed in a 7-day window in order to eliminate data granularity. Next, the data was converted into cases/100k inhabitants. All data analysis was conducted through in-house R scripts.

### 2.4 Natural Selection Analysis

Natural selection can manifest in directional, diversifying, purifying, and episodic selection. We evaluated these types of natural selection in Omicron genomes from Mexico. The Omicron SARS-COV-2 genome was annotated using RCoV19 (https://ngdc.cncb.ac.cn/ncov/?lang=en) (Gong et al. 2020). The directional selection search was performed with FEL (fixed effects likelihood), SLAC (single likelihood ancestor counting) (Delport et al. 2010), and Fast Unconstrained Bayesian AppRoximation (FUBAR), assuming that the selection pressure is constant across the phylogeny for each site (Murrell et al. 2013). Furthermore, the episodic diversifying selection was evaluated with the mixed-effects model (MEME) (Murrell et al. 2012).

#### Homoplasy test

We evaluated the sequences of Omicron (from Mexico) that have arisen independently several times (homoplasies). Homoplasies are likely candidates for adaptation due to convergent evolution, recombination, or errors during sequence data processing. Homoplasies were detected on phylogeny using HomoplasyFinder (Crispell, Balaz, y Gordon 2019). We collected all the Omicron sequences from Mexico and two sequences from Bostwana (NICD-N22418 and NICD-N22397) as reference genomes. Additionally, we used Snippy (https://github.com/tseemann/snippy) to detect single-nucleotide variations (SNVs) using the Omicron genome NICD-N22418 as reference genomes.

## 3. RESULTS

### 3.1. Phylogenetic evidence for multiple introductions Omicron and the human mobility facilitates its spread

On December 3, 2021, the first WGS-confirmed case of symptomatic COVID-19 caused by Omicron (B.1.1.519) was reported by the Instituto Nacional de Diagnóstico y Referencia Epidemiológica (InDRE), corresponding to a patient from South Africa that arrived at Mexico City on 16 November 2021. Since then and up to December 27, 2021, 254 genomes were sequenced in Mexico, corresponding to the states of Baja California (*n=1*), Chiapas (*n=1*), Guerrero (*n=1*), Hidalgo (*n=1*), Mexico City (*n =159*), Oaxaca (*n=1*), Puebla (*n=4*), Quintana Roo (*n =18*), Sinaloa (*n=2*), State of Mexico (*n=35*), Tabasco (*n=12*), Tamaulipas (*n=5*), Veracruz (*n=1*) and Yucatán (*n=13*). Interestingly, most of the Omicron sequences come from Mexico City and the State of Mexico, probably due to most international flights arriving in Mexico through the country’s capital. However, other states with international airports as well as important tourist destinations such as Baja California Sur, Quintana Roo, Yucatán, and Sinaloa also reported Omicron sequences and increases in cases in the first days of December (**Figure S1**).

To determine Omicron introductions to Mexico, we constructed a phylogenetic tree using 244 Omicron genome sequences from Mexico and 1903 foreign sequences deposited in GISAID. First, we inferred phylogeny for said sequences, to trace the movement of Omicron in Mexico through time and space. First, we note that the sequences of the Mexican genomes do not form a single clade but rather are scattered throughout the tree, consistent with multiple introductions to the country; In Figure A, the green branches correspond to the Mexican sequences, while the gray branches correspond to international sequences.

Through a discrete phylogeographic analysis following the Dellicour pipeline, introductions were evaluated (Dellicour, Durkin, et al. 2021). For this purpose, we used a time-scaled phylogenetic tree, and subsequently, introduction events were identified in the MCC tree with an associated HPD interval by reporting the number of introduction events. The analysis detected a minimum number of 173 introduction events of the Omicron lineage (95% HPD interval = [162.925-176.025]) identified from the phylogenetic analysis of 244 samples collected in Mexico (**Figure 1A**). The discrete phylogeographic analysis in BEAST show that the majority of introductions were represented by singletons (80%) and formed three small monophyletic clades composed of at least ten sequences from Mexico, corresponding to the center of Mexico, where the virus possibly initially spread throughout the country, driven by the mobility of people. However, the highest peak of Omicron in Mexico was reached by mid-January 2022, which may have had a more significant number of introductions.

**Figure 1.**
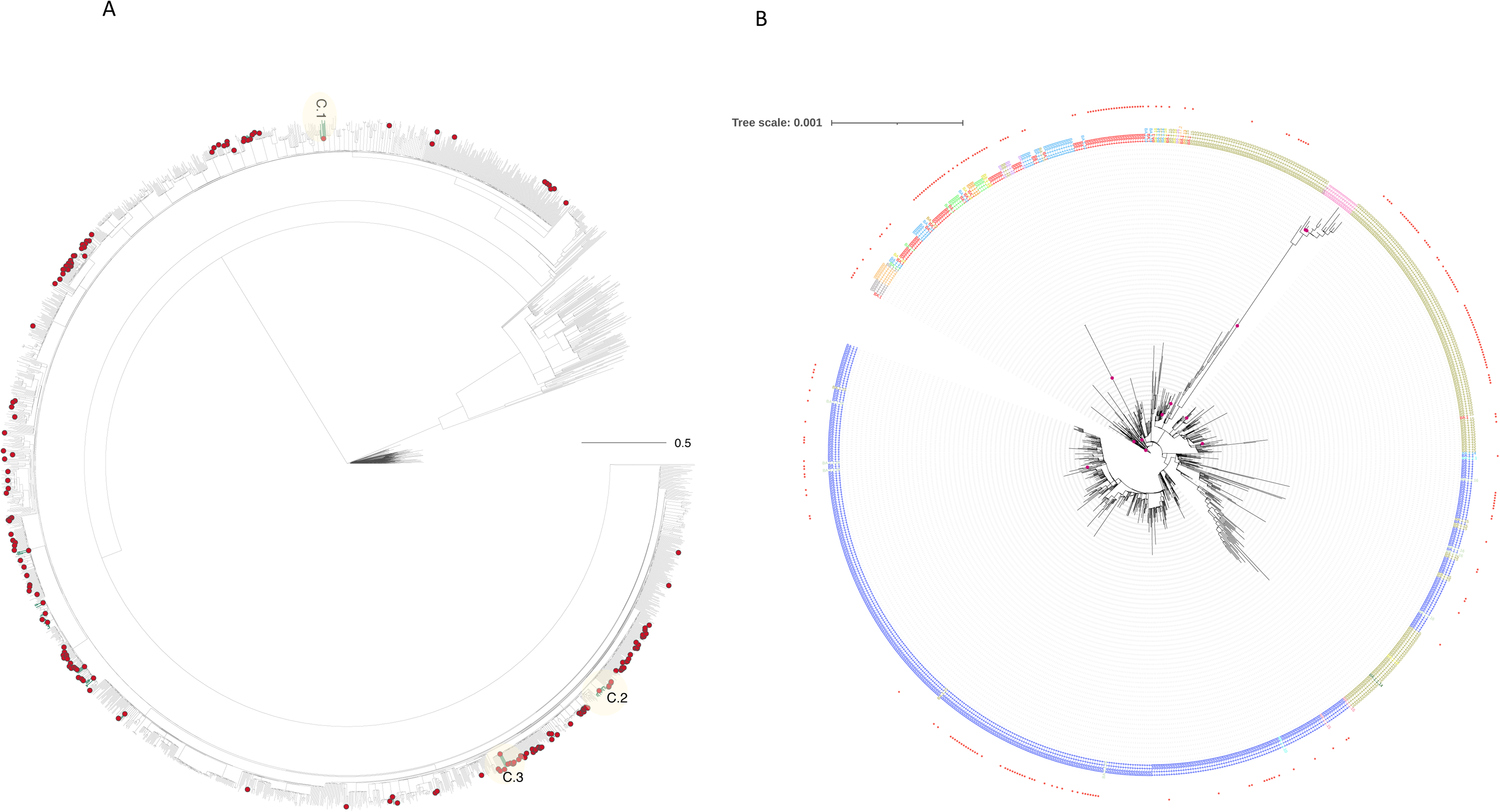
Phylogeography and Phylogenomics analysis Omicron. A) Time-scaled analysis of Omicron showing the 173 introduction events to Mexico, shown at the nodes in red color, and branches color in green represent genomes sampled from Mexico. In contrast, the branches in gray correspond to samples from other places. B) Maximum likelihood phylogeny of Omicron Mexican lineages, tips are colored by lineage.

The PastML analysis predicted Botswana as the location of the tree root of Omicron, suggesting that the Omicron variant origin might have been in southern African. Four main clusters emerged from the Botswana cluster; the oldest clusters were located in Hong Kong, India, Australia, and South Africa (**Figure S2**), which could indicate that the first transmissions and spreads of the virus were possibly in those locations. After the Botswana cluster, there is the cluster of South Africa, which is linked to one of India, from which a small cluster of Mexico emerges; this result suggests that the first independent introductions of Omicron to Mexico (four sequences) could have come from South Africa or India.

On the other hand, another small cluster from Mexico is connected to the Singapore cluster, which comes from a large group from Hong Kong (**Figure S2**). These results indicate that the introduction of the virus to Mexico could have been through different locations in Africa, India, and Asia. This analysis showed that the sequences of these regions share similarities with the first sequences of Mexico, and possibly, this was the spread of Omicron in the world towards Mexico.

As mentioned above, the peak of omicron was reached in mid-January 2022. A maximum-likelihood phylogenetic tree was built on understanding the distribution of Omicron during the increase in cases in the context of the first circulating genomes in Mexico. We found multiple introductions of different Omicron sublineages, which are marked with a red star in the phylogeny **(Figure 1B)**.

The first genomes that circulated during the months of November and December belong to the Omicron sub-lineages, a total of 244 genomes. **BA.1** 23.3% (n=57), **BA.1.1** 39% (n=97), **BA.1.1. 10** 0.4% (n=1), **BA.1.13** 0.4% (n=1), **BA.1.15** 29.91% (n=73), **BA.1.17** 1.6% (n=4), **BA.1.17.2** 0.4% (n=1), **BA.1.18** 0.81% (n=2), and **BA.1.7** 4.9% (n=12).

These lineages were identified for the first time in Mexico City. For example, **BA.1** on 16/11/2021 imported from South Africa; BA.**1.1** on 19/11/2021; BA.**1.1.10** on 21/12/2021. **BA.1.17** on 18/12/2021, BA.**1.17.2** on 16/12/2021, and **BA.1.18** on 10/12/2021. While BA.**1.15** was first identified on 03/12/2021 in Tamaulipas. Therefore, most of the introductions of the lineages are from the center of the country, and these introductions spread to the rest of Mexico.

In the tree, the first genomes are distributed within the clades of the omicron sub-lineages, indicating that these first introductions gave rise to the spread of the variant throughout the country. However, not all introductory lineages reached a high frequency, such as BA.1.7, BA.18, BA.1.17.2. Otherwise, lineages BA.1, BA.1.1, and BA.1.15 were the introductions that dominated and increased their frequency during the Omicron peak, which correspond to the largest clades in the phylogenetic tree (**Figure 1B**).

Interestingly, lineage BA.1.1 is subdivided into two large clades (clades in color Blue), followed by lineage BA.1.5 distributed in three clades (Green), followed by BA.1 (red), the most ancestral lineage in the tree. In contrast, the other lineages are found in a lesser proportion (**Figure 1B**).

We also investigated whether the introduction and spread of the Omicron variant could be due, in part, to trends of increased population mobility (**Figure 2**). We obtained human mobility data and divided the country into seven regions NE, NW, CN, CS, W, SE, and S. First, we found a mobility peak in December 2021 (**Figure 2-A**), involving all Mexican regions. The mobility increase observed in December 2021 is the highest observed in the Omicron selection period. Second, in **Figure 2-B** shows the increase in reported cases in January 2022. Next, in **Figure 2-C**, we analyze the correlation between increased mobility resulting in future cases. On average, it took six weeks for the correlation between the two variables to start showing a stronger correlation (except in the Northwest region). To explain the odd behavior of the Northwest region, in **Figure S3** the mobility of the states of Baja California, Chihuahua, Durango, and Sonora can be seen to increase earlier. Therefore, the earliest of these trends caused the correlation of this region to be different than the others. In **Figure 2-D**, The Omicron lineage peaked in mid-January 2022, where the B.1.1 lineage predominated throughout the country, followed by the BA.1.15 and BA.1 lineages. Therefore, the increase in cases is mainly associated with the BA.1.1 lineage. In addition, different sub-lineages of Omicron have been established, which have not increased in frequency, and perhaps many have become extinct.

**Figure 2.**
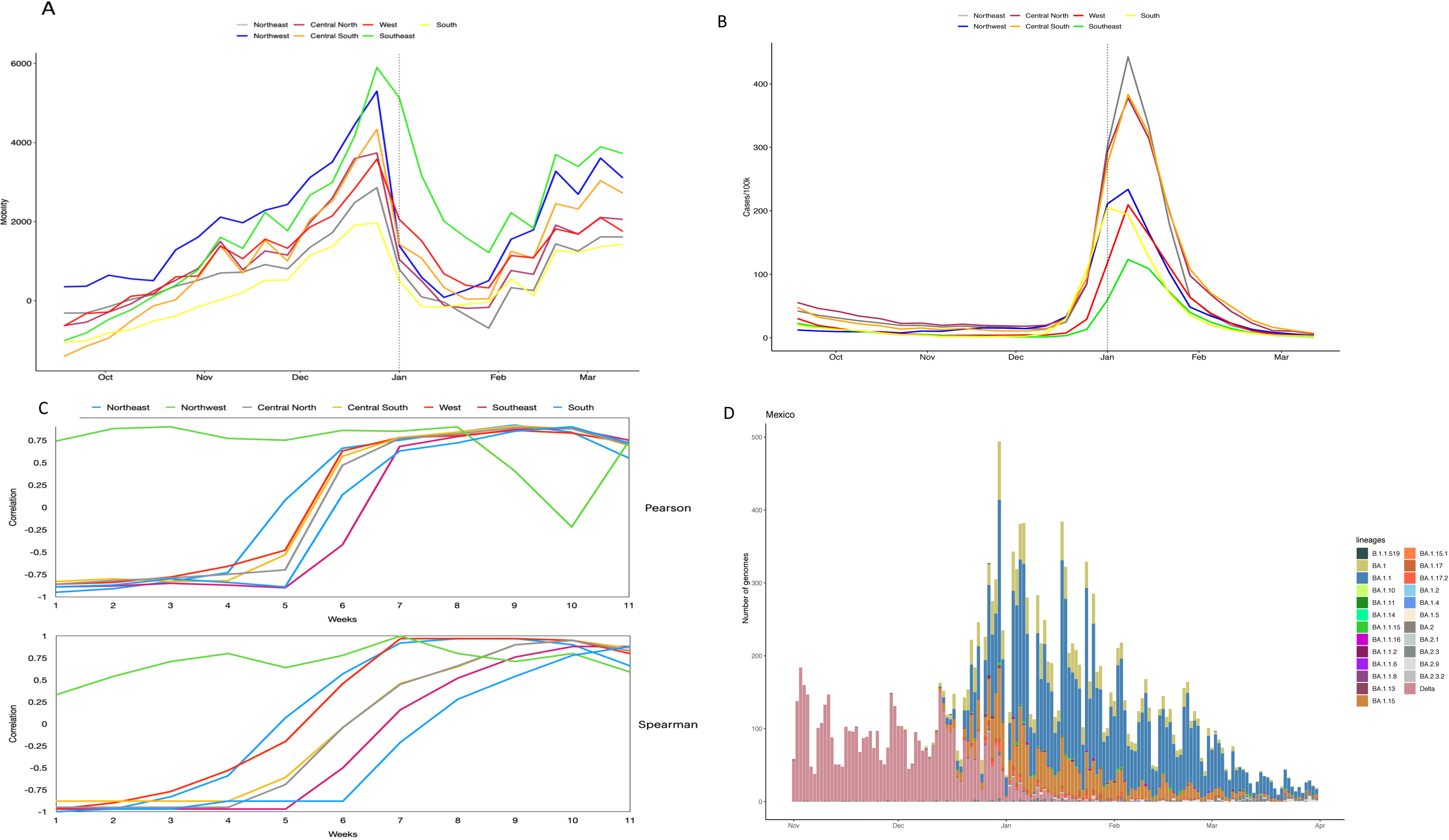
Mobility. A) Google mobility trends (retail, recreation, grocery, pharmacy, parks, public transport, workplace, and residential) of each Mexican region are computed from 2021-09-18 to 2022-03-12. Mobility levels above 0 indicate a trend above the baseline. The baseline was a period of observation from before the pandemic (the baseline day is the median value from the 5 week period Jan 3 – Feb 6, 2020. B) Daily COVID cases are computed in each Mexican region from 2021-09-18 to 2022-03-12. C) Pearson and Spearman correlations between COVID cases and mobility are computed. In order to analyze how long it takes for a spike in mobility to result in more cases, the cases were assessed in *n* weeks in the future (after the mobility spike). D) Number of SARS-CoV-2 genomes obtained at the national level. The BA.1.1 lineage was the most abundant in January and February. BA.2 in March, it seems its frequency begins to increase.

As mentioned above, the peak of mobility increased in December 2021 in all regions of Mexico; therefore, we also evaluated the prevalence and spread of omicron sub-lineages over time in the seven regions (**Figure 3**). Initially, in November, the prevalence of the Delta variant throughout the country was 99%. However, in December, there was an increase in mobility, and most of the cases this month in the Central and South region seem to be related to lineages BA.1 (∼10.80%), BA.1.1 (∼74.3%), and BA.1.15 (∼11.25%). Whereas some states are located on the US-Mexico border (Baja California, Chihuahua, Coahuila). On December NE and NW had a higher prevalence of Delta (70%) than the Central-South and South states that presented a 60% prevalence of Omicron sublineages. Subsequently, in January in all regions, Omicron was already dominant with the three sub-lineages BA.1, BA.1.1, and BA.1.1.15. Additionally, there was already a diversity of Omicron sub-lineages such as BA.2 (first time detected on 9/01/2022 in Jalisco), BA.1.1.10, BA.1.17, BA.1.17.2, BA.1.18 and BA.1.15. By February, BA.1.1 was already dominant over the other sub-lineages in all regions, while BA.2 in March began to increase in frequency, especially in the Central South and South regions.

**Figure 3.**
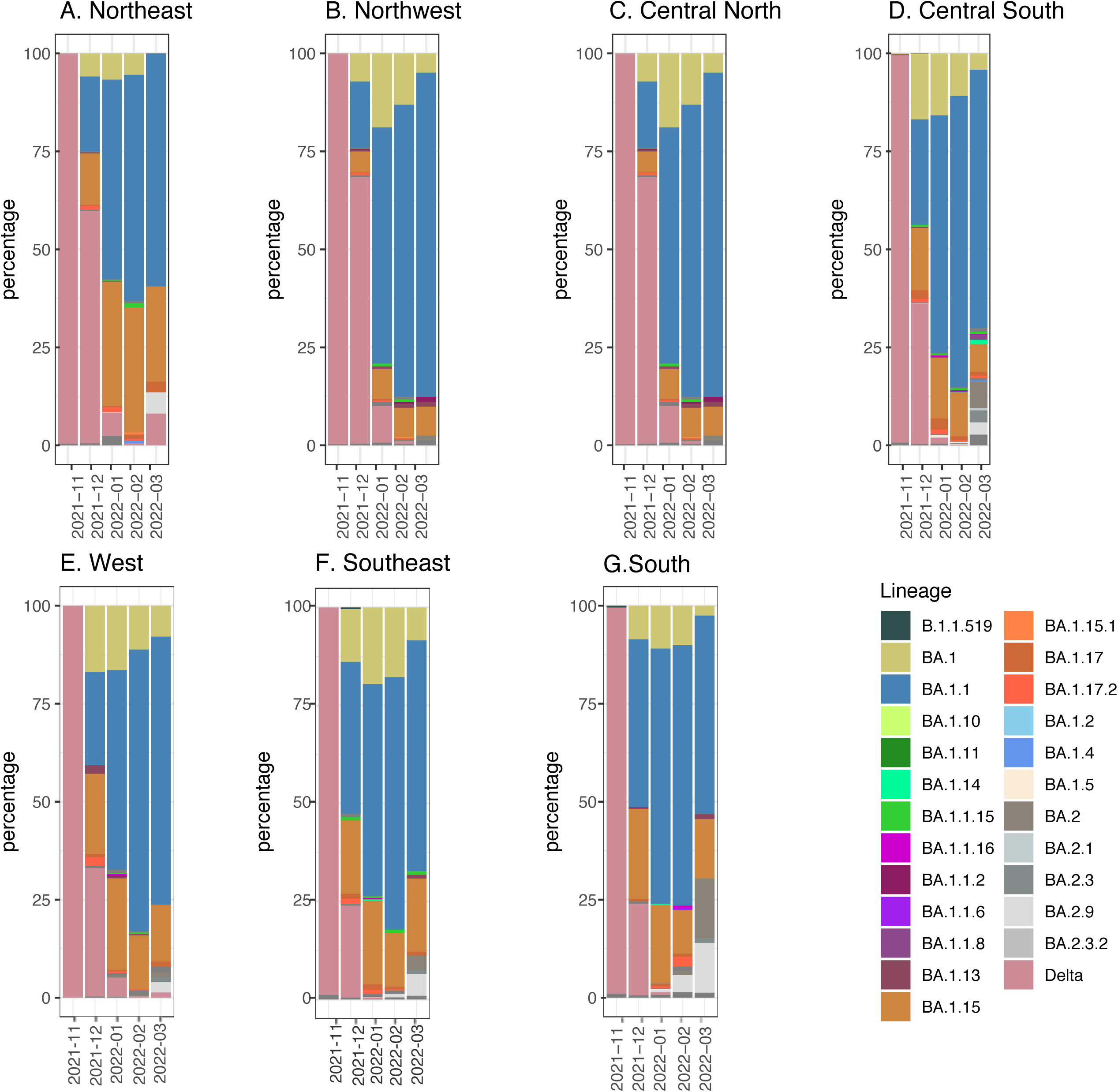
Distribution of Omicron lineages in Mexico between November 2021 and April 2022. (A), Northeast (B), Central North (C), Central South (D), West (E), Southeast (F), and South (G). The lineages B.1.1, B.1, and B.1.15 were predominant in all regions in January.

### 3.2 Analysis of natural selection of Omicron

We then searched for evidence of positive selection in Omicron genomes isolated from Mexico with different algorithms focused on a codon-based phylogenetic framework by calculating the ratio of non-synonymous (dN) to synonymous (dS) substitutions per coding sequence site (dN/dS). The evaluation of selection throughout the genome revealed that sites are highly conserved and show no evidence of episodic and directional selection. There was no evidence of an excess of non-synonymous mutations (dN/dS>1), representing potential purifying selection pressure, except for the gene encoding Spike. For the gene that codes for Spike protein, positive diversifying selection was detected on five codon sites, and non-synonymous changes in only five (dN/dS>1) through the FUBAR method, which assumes that the selection pressure for each site is constant along the entire phylogeny with a Bayesian algorithm to infer rates (dN/dS) (Murrell et al. 2013). Thus, genes were subjected to a distinct natural selection strength.

The sites under positive natural selection found in the Spike gene were; L371S, R346K, N417K, E484A, S496G, and N1098D (**Figure 4**). The L371S mutation (on Spike) is within the receptor-binding domain (RBD, residues numbered 319–541). The other observed mutations, R346K, N417K, E484A, and S496G are also found in the RBD. Mutation R346K described in the Mu variant can alter the interactions with monoclonal antibodies (Fratev 2022). Mutation E484A has been shown to confer up to 10-fold greater resistance to vaccinee sera (Wang et al. 2021, 1). The mutations L371S and S496G by neutral polar amino acid probably produce no difference in the receptor’s binding affinity (Rath, Padhi, y Mandal 2022). Finally, the N1098D mutation occurs in the spike protein’s S2 subunit (amino acid residues 686–1273). These changes produced in the Spike protein by charged polar amino acids could have significant repercussions, favoring a better interaction with the receptor. Interestingly, these mutations may be local adaptations of the virus to the Mexican population.

**Figure 4.**
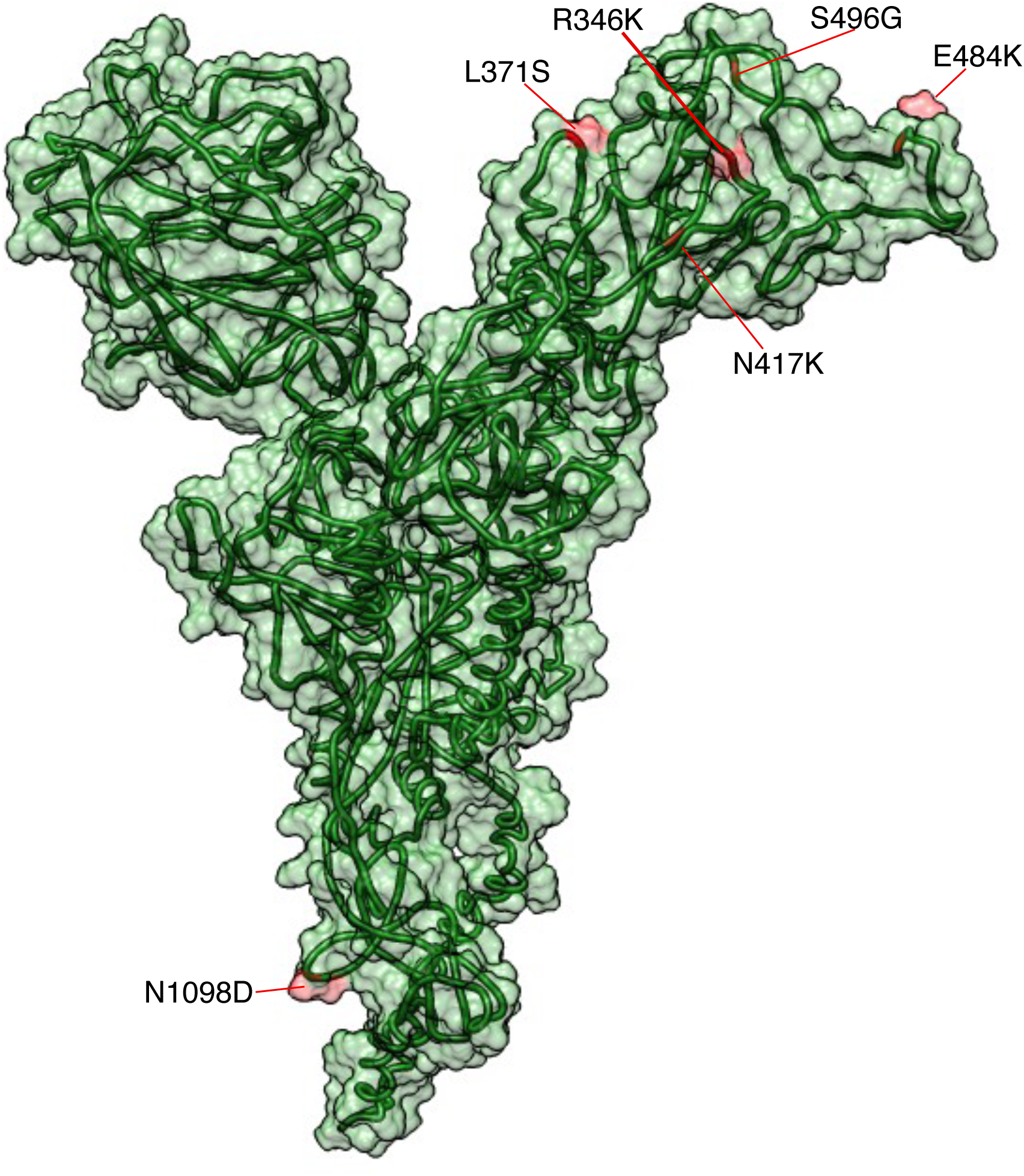
Spike protein model of Omicron SARS-COV. The sites that will evolve under positive selection according to FUBAR method are shown in red color.

### 3.3 Detection of Homoplasies

Homoplasies can be produced by base-pair substitutions or by recombination and can emerge naturally in the context of neutral evolution or positive natural selection. In order to detect homoplasies in the Omicron sequences in Mexico, we used HomoplasyFinder [24]. We used 244 Omicron (Mexico) sequences to construct a phylogeny, adding two South African genomes, NICD-N22418 and NICD-N22397, as the reference genomes. We identified homoplasies at 25 different positions (**Figure S4**), corresponding to 24.5% of the 102 polymorphic sites identified. Of these, 18 were in the ORF1a nsp11 gene. In addition, the homoplasy at position 15854 was in RNA polymerase, and the rest of the homoplasies (at positions 1694 to 9870) were in the non-structural proteins (nsp). The homoplasies detected tend to change for thymine or cytosine (**Supplementary table S1**).

On the other hand, using SNIPPY (using NICD-N22418 and NICD-N22397 as reference genomes), we found that the homoplasies at positions 1694, 2205, 5250, 5465 can be directly observed from the Botswana genomes, suggesting that the rest of the homoplasies could be generated from the descendants of the internal nodes.

## 4. DISCUSSION

In this study, we performed a phylogenetic and phylogeographic analysis to infer Omicron introductions in Mexico and how mobility might affect virus spread following introductions. Subsequently, we analyzed the adaptive evolutionary processes in local genomes during the first month of circulation of Omicron in Mexico. According to the data obtained from the genomes collected, the earliest detected genomes come mainly from Mexico City, the State of Mexico, Quintana Roo, and Yucatán, of which the first of the reported cases was imported from South Africa. However, most of the introduction cases in Mexico were singletons, which indicates multiple independent introductions (173 independent introductions). These results are like those reported during the first epidemic wave in New York City by Dellicour et al., 2021; discrete phylogeographic analysis found 116 independent introductions in the city and relatively small clades. These results show the heterogeneity of the virus to establish local transmission successfully (Dellicour, Hong, et al. 2021). The multiple introductions of variants to Mexico have already been explored in other lineages, such as B.1.1.519 (Taboada et al. 2021) and Alpha (Zárate et al. 2022). For example, in the Alpha lineage, the introductions probably came from the United States, contributing to the spread of the Alpha lineage from the north to the south of the country (Zárate et al. 2022)

However, in the case of Omicron, the increase in cases was in Mexico City, probably due to the arrival of international flights. As inferred from the PastML phylogeographic analysis, the Mexican clusters came from South Africa and India. The other introductions came from Hong Kong or Singapore, and this lineage arose from South Africa; Therefore, we argue that human mobility is a fundamental factor in introducing variants to a country. South Africa *per se* is an important airport hub in the continent (Newfarmer, Page, y Tarp 2018). An example of that is mobility data that shows that there have been approximately 93 thousand trips from South Africa to other 30 non-African regions by November 2021. Moreover, by the time the WHO had declared Omicron as a VoC, the probability of importation of the new variant exceeded 50% in North America, Europe, Middle East, and Oceania (Bai et al. 2021). Therefore, people’s inbound through international mobility is likely to be the introductory event of new variants and viruses.

From the moment a novel variant arrives in a country, internal mobility is among the factors that will facilitate its local transmission (Fakir y Bharati 2021; Oka, Wei, y Zhu 2021). We show that by observing a mobility increase and correlating it with the number of cases. In the regions where the first introductions of Omicron likely happened, i.e., Southwest, Northwest, and Central South, we observed that higher mobility was associated with higher cases six weeks after. On top of that, we argue that the mobility trends of a population follow a seasonal trend, as reported in (Hoogeveen, Kroes, y Hoogeveen 2022), since our data depicts a movement peak in the end-of-year season. Next, we show that the increase in cases reported in early January 2022 was due to the variants BA.1.1, BA.1, and BA.15, and it seems that there is no regionalization of the sub-lineages. And, the local transmission chains could reflect the higher internal mobility observed in the Southwest and Central South of the country; this has been observed in the Delta variant in Mexico […].

Once knowing the introductions of omicron to Mexico, we evaluated the adaptive changes in the genomes of Mexico. Since natural selection is one of the main evolutionary forces that generate diversity and show signs of adaptation of an organism, we evaluate the diversifying directional selection. We use FUBAR, which shows better performance on SLAC and FEL (Delport et al. 2010; Murrell et al. 2013). The results revealed six sites under positive selection on Spike protein (dN/dS>1). The residues under positive selection are located in RBD on spike protein surrounding the receptor-binding site (R346K, L371S, K417N, E484A, S496G, and N1098D) associated manly with evasion of antibodies. For example, the R346K amino acid change, previously detected in lineage Mu (B.1.621) (Laiton-Donato et al. 2021, 2) is associated with increased resistance to convalescent plasma therapy, and less neutralization of antibodies(Liu et al. 2022). The substitution R346K was previously reported in genomes from Mexico City with a prevalence of 40% in the capital and according to phylogenetic analyses, the sequences from Mexico City form a monophyletic clade (Cedro-Tanda et al. 2022). Genomes with the R346K mutation now designated as BA.1.1 sub-lineage (Mohandas et al. 2022) which predominated in Mexico and spread throughout the country.

The amino acid substitution L371S has been associated with a significantly increased binding affinity to human ACE2 (Kimura et al. 2022). The amino acid substitution K417N has been previously reported in the Alpha and Gamma lineages, which facilitates the escape of monoclonal antibodies (bamlanivimab/LY-CoV555) (Starr et al. 2021, 016) and the escape of vaccine sera such as BNT162b2 (Hoffmann et al. 2021). The amino acid substitution E484A is under selection in genomes in Mexico; this substitution was previously E484K in the Beta, Gamma, and Mu variants related to the escape to neutralization of vaccines. Currently, mutation E484A is present in Omicron, weakening the mAbs binding to escape the immune response (Shah y Woo 2022). The S496G amino acid change is related to evasion of mAbs binding (Kannan et al. 2022).

In general, the sites under selection in the protein S of the Omicron variants in Mexico are associated with the evasion of antibodies. In addition, several recent studies have proven that natural selection is evident in the Spike a glycoprotein (Chaw et al. 2020; Tang et al. 2020). Changes in the amino acid sequence of RBD can dramatically impact the binding affinity of Spike for the ACE2 receptor, in addition to causing infectivity and transmission of SARS-CoV-2.

The methods used to detect natural selection determined sites under selection in Spike and not in other Omicron genome regions. However, ORF1ab, ORF3a, and ORF8 proteins have been reported to have sites that evolve under positive natural selection (Velazquez-Salinas et al. 2020). Many of the mutations can occur repeatedly and independently throughout the genome several times (homoplasies), which produce non-synonymous changes at the protein level, are candidates for possible adaptation due to positive natural selection. Therefore, we evaluated the presence of homoplasies, which are recurrent mutations in the genomes of Mexico; we found that most of the changes were associated with the ORF1a nsp11 gene, RNA polymerase, and non-structural proteins (nsp). The recurrent mutation signal in Orf1a, which encodes the nonstructural proteins Nsp11 and Nsp13, has been previously reported (van Dorp et al. 2020, 2). In our analysis, the Nsp11 was the recurrent gene with homoplasies.

## 5. CONCLUSIONS

The analyses presented here show multiple introductions of omicron that took place in different regions of central-southern Mexico, then spreading to the rest of the country during the first months of its circulation. We have also gathered evidence to show that higher levels of mobility may have facilitated the spread of the variant to the outer regions of the country. Here, we show that the increased mobility at the end of the year resulted in a peak of Omicron cases; on average, it started being observed six weeks later. The identification of introductions is essential to determine the spread of the virus in a country. It also highlights the presence of diversifying natural selection in the Spike protein and the presence of homoplasies mutations in non-structural proteins, which may be possible candidates for adaptation of the virus to the Mexican population. It is crucial to identify possible adaptation signatures in SARS-CoV-2 for the continued development of vaccines and treatments.

## Data Availability

All data produced in the present work are contained in the manuscript

## Acknowledgments

We thank to Consorcio Mexicano de Vigilancia Genómica (CoviGen).

We thank the submitting laboratories for generating the genetic sequence and metadata and sharing it through the GISAID initiative, on which this research is based.

## Authors contributions

HGCS conceived the initial study. HGCS, GLL, JTF and LMC designed the experiments. HGCS, GLL, and GSM performed analysis. HGCS, GLL, GMS, JTF, and LMC have contributed to the interpretation and discussion of the results. HCGS and GLL wrote the first manuscript draft with editing from GMS, JTF, and LMC. The final manuscript was approved by all authors.

## Figure Supplementary

**Figure S1. Number of cases per 100,000 population**.

The number of cases per 1000 inhabitants in the first months of the introduction of omicron is shown in each state of Mexico. The states of Baja California Sur, Mexico City, State of Mexico, Quintana Roo, and Yucatan.

**Figure S2. Ancestral-state reconstruction using maximum likelihood inferred with PastML**.

Colors correspond to different countries. Numbers inside (or next to) the circles indicate the number of strains assigned to the specific node, and the size of the circles is proportional to the number of tips.

**Figure S3. Mobility of Northwest region**.

The states composing the Northwest region were isolated in terms of their mobility trends in order to explain the odd behavior of the mobility *vs*. cases correlation reported in the Northwest region by Figure 2-C. The mobility index reported is the result of the mobility categories Google displays (i.e., retail, recreation, grocery, pharmacy, parks, public transport, workplace, and residential). The baseline indicates how mobility used to be in the timespan of Jan 3 – Feb 6, 2020.

**Figure S4. Homoplasies in Omicron from Mexico**.

A phylogenetic tree was reconstructed using 244 Omicron genome sequences. The colored right columns represent the 25 homoplasies identified. The nucleotides associated with these positions in each sequence are plotted and colored according to their type (Adenine=red, Cytosine=blue, Guanine=cyan, Thymine=orange, and no information for the nucleotide (Ns) = white). The tree’s base is in the NICD-N22418 and NICD-N22397 as reference genomes. Homoplasies are indicated in the internal nodes (Mexican sequences).

**Supplementary Table S1**

Supplementary Table 1. The number of changes observed (multiple alleles present) for each of sites on the phylogeny.

